# Higher Family Income and Clinical Dental Outcomes of Minority Children: A Cross-Sectional Study

**DOI:** 10.1101/2023.05.08.23289674

**Authors:** David Okuji, Rolina Luo, Tianqi Wei, Myeonggyun Lee

**Author notes:** **Corresponding Author:** David Okuji, DDS, MBA, MS, NYU Langone Hospitals, 5800 Third Avenue, 3^rd^ Floor, Brooklyn, NY 11220.

## Abstract

**Purpose:** This study utilized Minorities’ Diminished Return theory and aimed to learn if minority children gain less protective effect from higher family income compared to White children for dental caries and multiple oral health problems.

**Methods:** This study was designed cross-sectionally from 21,599 subject-responses to the 2017 National Survey of Children’s Health. Specific outcomes included parental-reported dental caries and multiple oral health problems (e.g., toothaches, bleeding gums, and/or caries) compared across Hispanic and non-Hispanic White, Black, Asian, and Multi-race children. Logistic regression models estimated the effects of race/ethnicity on each outcome, with adjustments for child sex, parental education, child age, and income-to-needs ratio.

**Results:** The findings showed when all racial/ethnic groups increased family income and socioeconomic status, there were no statistically significant differences between all racial/ethnic groups for the parental-reported dental caries and multiple oral health problems.

**Conclusions:** This study’s findings show either 1) Minorities’ Diminished Return theory, which postulates that structural racism negatively impacts health gains from higher socioeconomic status for minority children, does not apply to children’s dental caries and multiple oral health problems or 2) the biologic load of the caries disease process supersedes the protective health gain effects from increased family income, expected by Minorities’ Diminished Return theory. Regardless, dentists and policymakers must support best practices and policies which benefit entire populations of children, through widespread disease prevention techniques, access-to-care, and utilization-of-care programs to decrease levels of childhood caries and other oral health problems.

## Introduction

In 2020, the American Medical Association (**AMA**) issued a press release stating, the “AMA Board of Trustees today pledged action to confront systemic racism” and “recognizes that racism in its systemic, structural, institutional, and interpersonal forms is an urgent threat to public health, the advancement of health equity, and a barrier to excellence in the delivery of medical care.”^1^ “Systemic racism” encompasses whole systems which include social determinants of health such as political, legal, economic, healthcare, educational, and criminal justice system and includes the structures that uphold the systems.^2^ “Structural racism” focuses on the role of structures such as, laws, policies, institutional practices, and entrenched standards that support the system.^3^

From a systemic perspective, despite the growing number of healthcare providers, improvement in medical and oral healthcare, and breakthroughs in medications; racial/ethnic minority groups of children persistently experience multiple disparities across different health fields and have substandard health conditions compared to privileged groups of children.^4-5^ Examples of health disparities experienced by low-income, minority children include asthma, mental disorder, heart diseases, and kidney diseases.^6-8^

Oral health disparities have not been studied as extensively as medical health disparities, but poor oral health has significant influence on the overall body health of children and adults.^9^ The prevalence of untreated dental caries in African American and Caucasian children between ages 3-5 years old was 19% and 11%, respectively. The difference between the prevalence of untreated dental caries in African American and Caucasian teenagers of age 13 to 15 years old was even higher.^9^ Multiple studies have shown that oral health disparities for the African American community are prevalent and influenced by a multitude of factors, including sociocultural context, structure factors, and family experience.^9-10^ Lower income level and cultural perspective can further alter the behavior and perception of seeking treatments. It is essential that oral healthcare professionals understand how certain racial/ethnic groups have a higher incidence of oral diseases and a lower treatment success rate due to those oral health disparities.^11^

Minorities’ Diminished Return (**MDR**) theory is a social epidemiology construct which postulates minority populations receive lower increases in health gains compared to White populations when both minority and White families increase family income and socioeconomic status.^12-15^ MDR theory is based upon differential group vulnerability, which hypothesizes that equal resources result in unequal outcomes, with marginalized groups being systemically disadvantaged relative to the dominant group. MDR theory postulates that lower gains for health outcomes of marginalized groups are because their resources generate less tangible health gains than the privileged group.^16^ Although previous studies revealed the effect of MDR theory on unmet dental care need^12^ (**DCN**) and asthma^15^ between White and Black children, there is no evidence regarding the association between MDR theory and the clinical outcomes of parental-reported dental caries and other oral health problems, such as toothaches and/or bleeding gums, across other racial minority groups of children.

Therefore, the purpose of this study was to analyze the 2017 National Survey of Children’s Health (**NSCH**) data set^17^ and apply MDR theory to compare the effect of higher income-to-needs ratio, a proxy for socioeconomic status (**SES**), between minority families and White families for their parental-reported children’s dental caries and multiple oral health problem experiences. The null hypothesis was all children regardless of race/ethnicity would receive equal health gains for prevalence of parental-reported dental caries and multiple oral health problem experiences when income-to-needs ratio increased.

## Method

### Study Design and Setting

The present study was designed as an analytical, observational, cross-sectional model, which examined secondary data from the 2017 NSCH. The collected data related to the physical and mental health of American children from birth to 17 years old in the 50 states plus the District of Columbia. The study analyzed weighted, nationally representative data utilizing the U.S. Census Bureau racial/ethnic group classifications of Hispanic and Non-Hispanic White, Black, Asian, and Multi-race.^17^

### Study Size, Participants, and Data Sources

A total of 59,135 sample households in the 50 states plus the District of Columbia were initially screened to find eligible children based on age. From all the inclusion-eligible children, the caregivers of 21,599 children completed the topical questionnaire about their children’s physical and mental health with language availability in English and Spanish.^17^ Details of the NSCH subject recruitment process and inclusion criteria were accessed from the NSCH Codebooks.^17^

### Variables and Data Measurement

The current study’s outcomes of interest were defined as dichotomous variables indicating an answer of “yes” or “no” to the questions: 1) parental-reported dental caries (**DC**) measured with a “yes” response to the question, ‘Does this child have cavities or tooth decay?; and”, 2) parental-reported multiple oral health problems (**MOHP**) measured with a ‘yes’ response to the question, ‘Does this child have one or more oral health problems?’ (i.e., toothaches, bleeding gums, decayed teeth, or cavities)”.

Additional variables extracted from the NSCH data set included the following parameters: 1) child age as a continuous variable, 2) child sex as a binary variable of Male or Female, 3) child race and ethnicity defined by non-migrant Hispanic (**Hispanic**), non-Hispanic White (**White**), non-Hispanic Black (**Black**), non-Hispanic Asian (**Asian**), and non-Hispanic Multi-race/ethnicity (**Multi-race**) families, 4) parental education defined by “Less than high school,” “High school degree or General Education Diploma (**GED**),” “Some college or technical school,” and “College degree or higher” as an ordinal variable, and 5) income-to-needs ratio, a proxy for SES based on ordinal variables calculated as a continuous variable of the “family poverty ratio” from the NSCH data. The family poverty ratio was calculated as a percentage of the ratio of total family income and the family poverty threshold, based upon family size. Note that income-to-”needs” ratio does not imply “dental needs” and instead reflects adequacy of family income compared to family size.

### Statistical Methods and Quantitative Variables

For descriptive statistics, continuous variables were reported with mean and standard deviations and categorical variables were summarized with frequencies and percentages. The descriptive statistics were reported overall in the pooled data. The distributions of variables were compared across different race/ethnicity groups with the analysis of variance (**ANOVA**) test for continuous variables and the Chi-square test for categorical variables. The missing data were excluded in the analysis given their low frequency and the observed data with sampling weights.

The logistic regression models were fitted to estimate the effects of race/ethnicity on each outcome, with adjustments for child sex, parental education, child age, and income-to-needs ratio. For each outcome, this study considered two logistic models: Model 1) only outcomes and Model 2) outcomes with the interaction between race/ethnicity and income-to-needs ratio. To investigate the effects of each variable on outcomes within the race/ethnicity group, the analysis additionally conducted the logistic regression analysis, stratified by race/ethnicity.

All statistical tests were two-sided, with Wald type confidence intervals provided, and p<0.05 was considered statistically significant. All analyses were performed in R version 4.0.3 (The R Foundation for Statistical Computing, Vienna, Austria).^18^ Note that the NSCH data set utilized sampling weights for data analysis in the study to attain population-based estimates. To account for the NSCH weights, *‘survey’* R package was used for the analyses.

### Ethics

The NYU Grossman School of Medicine Institutional Review Board reviewed and determined this study (Protocol number 20-01537) did not qualify as human-subjects research.

## Results

Table 1 shows the descriptive analyses and statistically significant inferential associations. Descriptively, the sample population was mostly male (51.2 percent), with mean age of 8.6 years old, 49.6 percent of parents with their highest educational level of a college degree or higher, and mean income-to-needs ratio of 246.17 percent. Inferentially, associations were demonstrated with income-to-needs ratio and parental high school education. Black and Hispanic children had lower socioeconomic levels, reflected by lower income-to-needs ratio, and higher levels of parental high school or General Education Diploma as their highest education level. Among the outcomes, only MOHP was statistically significant, with Black (17.8 percent), Hispanic (16.8 percent), and Asian (17.4 percent) children having higher prevalence for MOHP. A post-hoc power analysis demonstrated 100% power (1-beta) with alpha set at 0.05.

**Table 1.**
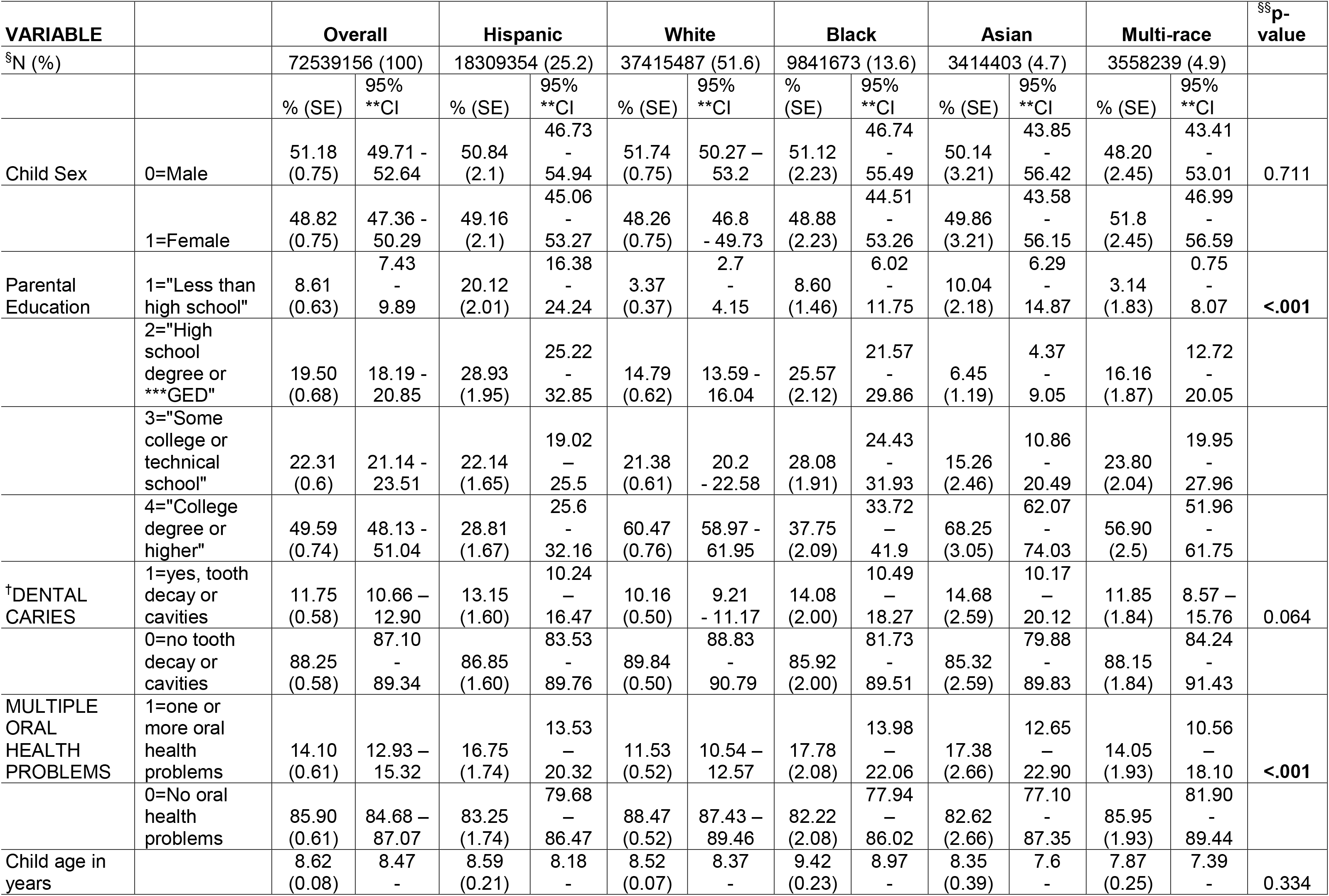
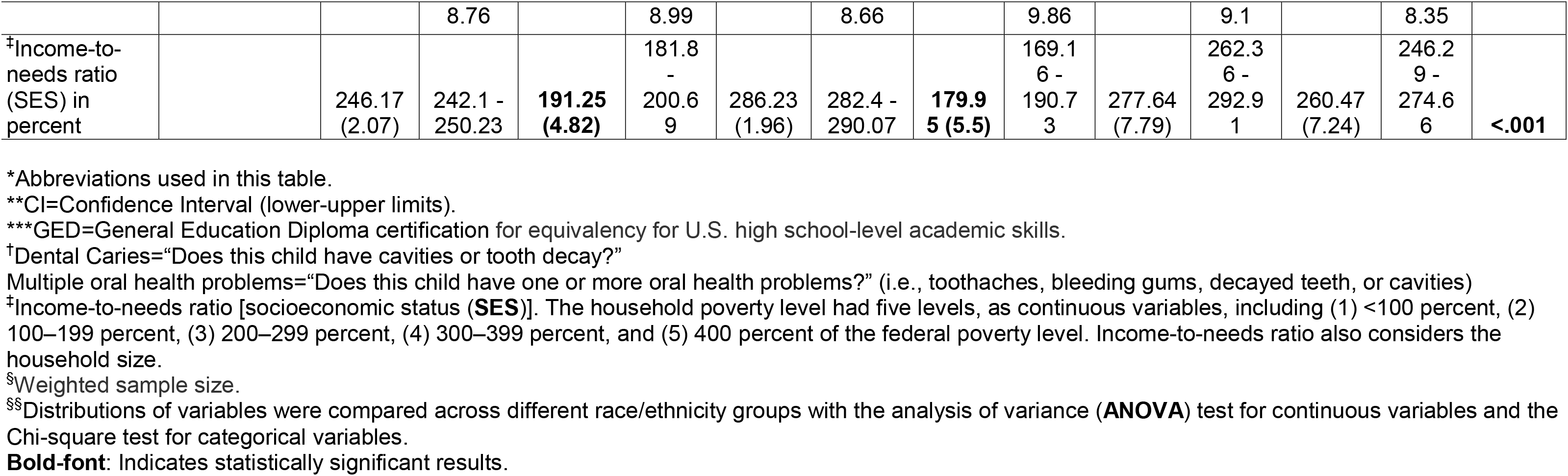
Patient demographics and outcome variables overall, by race*.

Table 2 presents the Model 1 summary of logistic regressions for DC and MOHP, which were estimated in the pooled sample. Model 1 only included the main outcomes. Table 3 presents the Model 2 summary of logistic regressions for DC and MOHP, which were estimated in the pooled sample. Model 2 added the race/ethnicity by income-to-needs ratio interaction. Both Table 2 and Table 3 utilized White race/ethnicity as the reference.

**Table 2.**
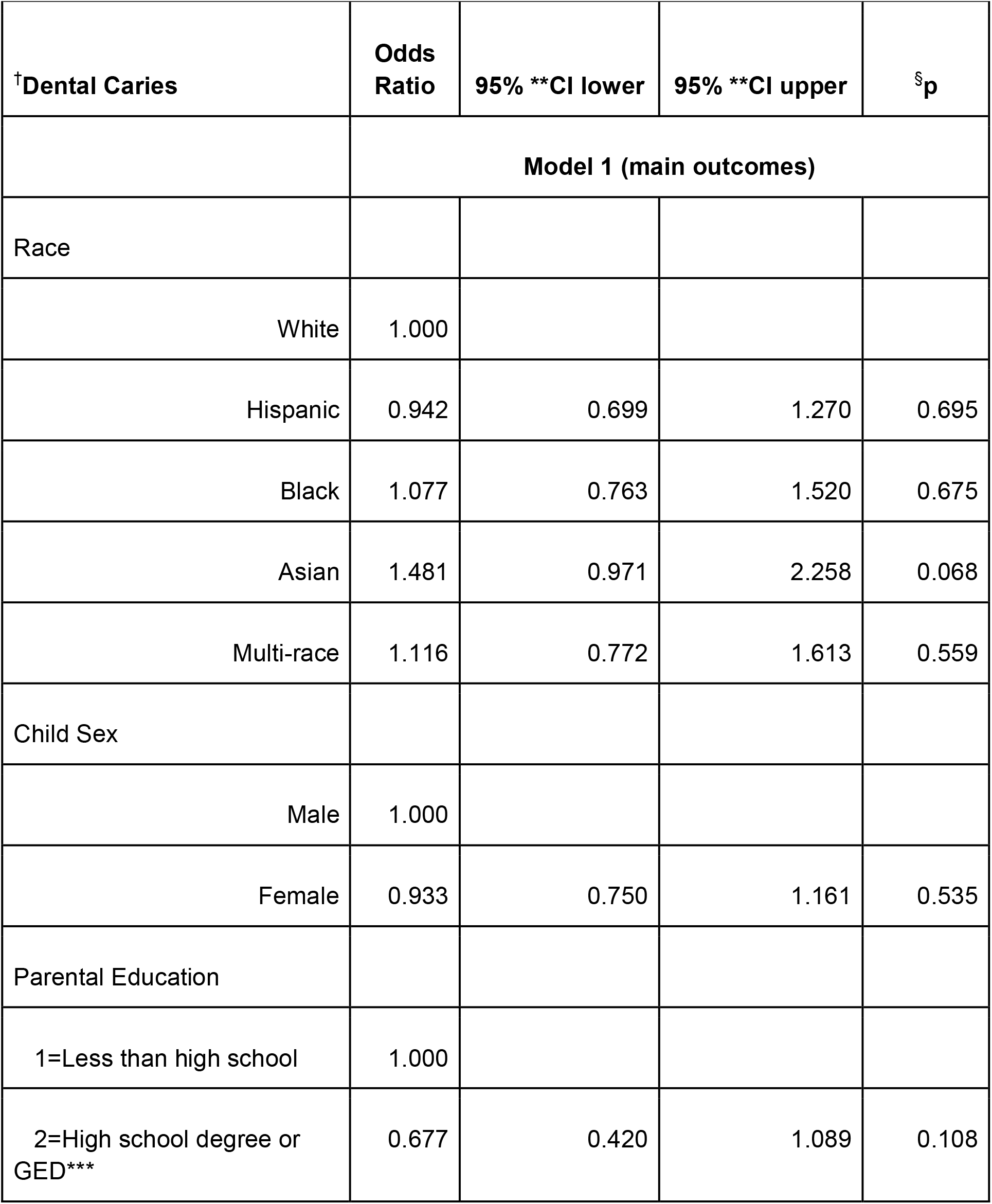
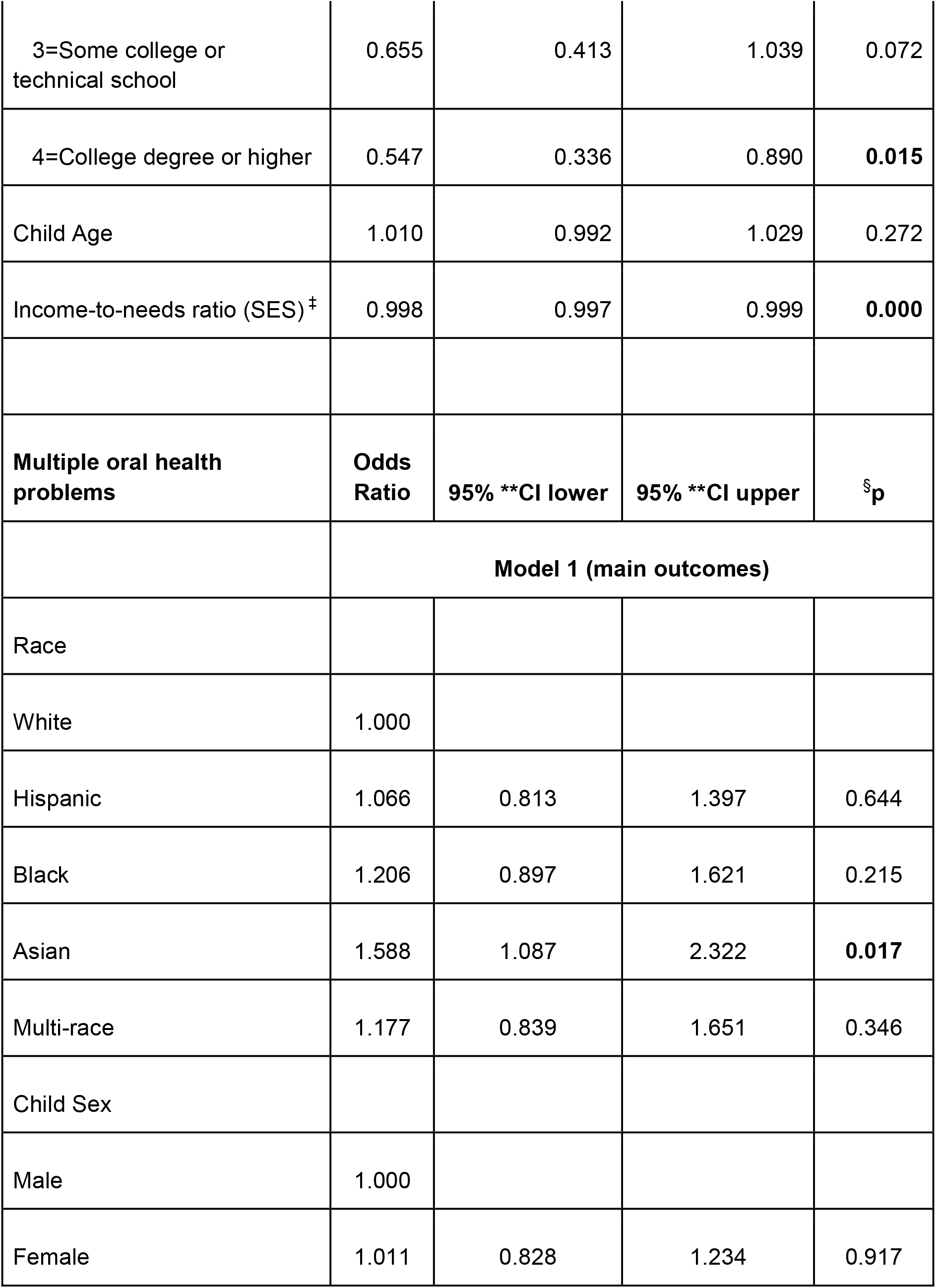
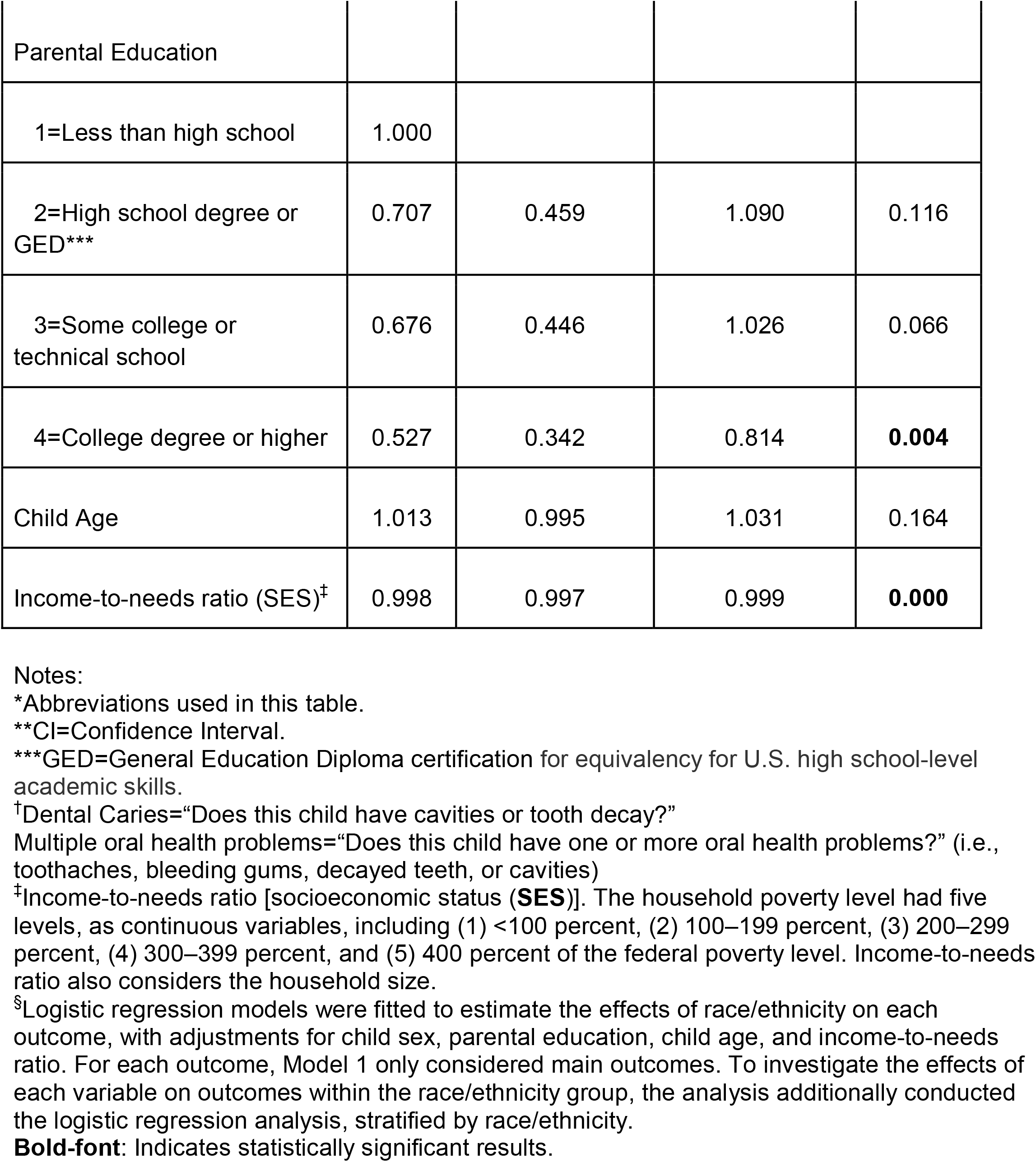
Pooled Sample Logistic Regressions for Access to dental care, Utilization of preventive dental care, and Utilization of any dental care; with Model 1 (main outcomes)*.

| <sup>†</sup>Dental Caries | Odds Ratio | 95% **CI lower | 95% **CI upper | <sup>§</sup>p |
| --- | --- | --- | --- | --- |
|  | **Model 1 (main outcomes)** |  |  |  |
| Race |  |  |  |  |
| White | 1.000 |  |  |  |
| Hispanic | 0.942 | 0.699 | 1.270 | 0.695 |
| Black | 1.077 | 0.763 | 1.520 | 0.675 |
| Asian | 1.481 | 0.971 | 2.258 | 0.068 |
| Multi-race | 1.116 | 0.772 | 1.613 | 0.559 |
| Child Sex |  |  |  |  |
| Male | 1.000 |  |  |  |
| Female | 0.933 | 0.750 | 1.161 | 0.535 |
| Parental Education |  |  |  |  |
| 1=Less than high school | 1.000 |  |  |  |
| 2=High school degree or GED*** | 0.677 | 0.420 | 1.089 | 0.108 |
| 3=Some college or technical school | 0.655 | 0.413 | 1.039 | 0.072 |
| 4=College degree or higher | 0.547 | 0.336 | 0.890 | **0.015** |
| Child Age | 1.010 | 0.992 | 1.029 | 0.272 |
| Income-to-needs ratio (SES)<sup>‡</sup> | 0.998 | 0.997 | 0.999 | **0.000** |
| **Multiple oral health problems** | **Odds Ratio** | **95% \*\*CI lower** | **95% \*\*CI upper** | **§p** |
|  | **Model 1 (main outcomes)** |  |  |  |
| Race |  |  |  |  |
| White | 1.000 |  |  |  |
| Hispanic | 1.066 | 0.813 | 1.397 | 0.644 |
| Black | 1.206 | 0.897 | 1.621 | 0.215 |
| Asian | 1.588 | 1.087 | 2.322 | **0.017** |
| Multi-race | 1.177 | 0.839 | 1.651 | 0.346 |
| Child Sex |  |  |  |  |
| Male | 1.000 |  |  |  |
| Female | 1.011 | 0.828 | 1.234 | 0.917 |
| Parental Education |  |  |  |  |
| 1=Less than high school | 1.000 |  |  |  |
| 2=High school degree or GED*** | 0.707 | 0.459 | 1.090 | 0.116 |
| 3=Some college or technical school | 0.676 | 0.446 | 1.026 | 0.066 |
| 4=College degree or higher | 0.527 | 0.342 | 0.814 | **0.004** |
| Child Age | 1.013 | 0.995 | 1.031 | 0.164 |
| Income-to-needs ratio (SES)<sup>‡</sup> | 0.998 | 0.997 | 0.999 | **0.000** |

Notes:
\*Abbreviations used in this table.
\*\*CI=Confidence Interval.
\*\*\*GED=General Education Diploma certification for equivalency for U.S. high school-level academic skills.
<sup>†</sup>Dental Caries="Does this child have cavities or tooth decay?"
Multiple oral health problems="Does this child have one or more oral health problems?" (i.e., toothaches, bleeding gums, decayed teeth, or cavities)
<sup>‡</sup>Income-to-needs ratio [socioeconomic status (**SES**)]. The household poverty level had five levels, as continuous variables, including (1) <100 percent, (2) 100–199 percent, (3) 200–299 percent, (4) 300–399 percent, and (5) 400 percent of the federal poverty level. Income-to-needs ratio also considers the household size.
<sup>§</sup>Logistic regression models were fitted to estimate the effects of race/ethnicity on each outcome, with adjustments for child sex, parental education, child age, and income-to-needs ratio. For each outcome, Model 1 only considered main outcomes. To investigate the effects of each variable on outcomes within the race/ethnicity group, the analysis additionally conducted the logistic regression analysis, stratified by race/ethnicity.
**Bold-font:** Indicates statistically significant results.

**Table 3.** Pooled Sample Logistic Regressions for Access to dental care, Utilization of preventive dental care, and Utilization of any dental care; with Model 2 (interaction between outcome of race and income-to-needs ratio)*.

| †Dental Caries | Odds Ratio | 95% **CI lower | 95% **CI upper | §p |
| --- | --- | --- | --- | --- |
|  | **Model 2 (race by income-to-needs ratio interaction)** |  |  |  |
| Race |  |  |  |  |
| White | 1.000 |  |  |  |
| Hispanic | 0.296 | 0.185 | 0.474 | **0.000** |
| Black | 1.050 | 0.583 | 1.892 | 0.871 |
| Asian | 0.992 | 0.546 | 1.803 | 0.978 |
| Multi-race | 1.907 | 0.757 | 4.801 | 0.171 |
| Child Sex |  |  |  |  |
| Male | 1.000 |  |  |  |
| Female | 0.936 | 0.752 | 1.163 | 0.549 |
| Parental Education |  |  |  |  |
| 1=Less than high school | 1.000 |  |  |  |
| 2=High school degree or GED*** | 0.687 | 0.424 | 1.115 | 0.128 |
| 3=Some college or technical school | 0.662 | 0.412 | 1.062 | 0.087 |
| 4=College degree or higher | 0.553 | 0.337 | 0.910 | **0.020** |
| Child Age | 1.010 | 0.992 | 1.029 | 0.281 |
| Income-to-needs ratio (SES)<sup>†</sup> | 0.998 | 0.997 | 0.999 | **0.000** |
| Income-to-needs ratio X Race |  |  |  |  |
| status:Hispanic | 0.999 | 0.997 | 1.002 | 0.645 |
| status:Black | 1.001 | 0.998 | 1.003 | 0.652 |
| status:Asian | 0.999 | 0.996 | 1.002 | 0.466 |
| status:Multi-race | 0.998 | 0.996 | 1.001 | 0.192 |
| **Multiple oral health problems** | **Odds Ratio** | **95% \*\*CI lower** | **95% \*\*CI upper** | **§p** |
|  | **Model 2 (race by income-to-needs ratio interaction)** |  |  |  |
| Race |  |  |  |  |
| White | 1.000 |  |  |  |
| Hispanic | 1.248 | 0.740 | 2.104 | 0.406 |
| Black | 1.239 | 0.740 | 2.073 | 0.416 |
| Asian | 2.497 | 1.117 | 5.583 | **0.026** |
| Multi-race | 1.539 | 0.764 | 3.101 | 0.228 |
| Child Sex |  |  |  |  |
| Male | 1.000 |  |  |  |
| Female | 1.012 | 0.829 | 1.236 | 0.904 |
| Parental Education |  |  |  |  |
| 1=Less than high school | 1.000 |  |  |  |
| 2=High school degree or GED*** | 1.627 | 1.066 | 2.483 | **0.024** |
| 3=Some college or technical school | 1.874 | 1.247 | 2.818 | **0.003** |
| 4=College degree or higher | 3.101 | 2.100 | 4.580 | **0.000** |
| Child Age | 1.013 | 0.995 | 1.031 | 0.172 |
| Income-to-needs ratio (SES)<sup>†</sup> | 0.998 | 0.997 | 0.999 | **0.000** |
| Income-to-needs ratio X Race |  |  |  |  |
| status:Hispanic | 0.999 | 0.997 | 1.001 | 0.477 |
| status:Black | 1.000 | 0.998 | 1.003 | 0.966 |
| status:Asian | 0.998 | 0.996 | 1.001 | 0.140 |
| status:Multi-race | 0.999 | 0.996 | 1.001 | 0.321 |

Notes:
\*Abbreviations used in this table.
\*\*CI=Confidence Interval.
\*\*\*GED=General Education Diploma certification for equivalency for U.S. high school-level academic skills.
†Dental Caries="Does this child have cavities or tooth decay?"
Multiple oral health problems="Does this child have one or more oral health problems?" (i.e., toothaches, bleeding gums, decayed teeth, or cavities)
‡Income-to-needs ratio [socioeconomic status (**SES**)]. The household poverty level had five levels, as continuous variables, including (1) <100 percent, (2) 100–199 percent, (3) 200–299 percent, (4) 300–399 percent, and (5) 400 percent of the federal poverty level. Income-to-needs ratio also considers the household size.
§Logistic regression models were fitted to estimate the effects of race/ethnicity on each outcome, with adjustments for child sex, parental education, child age, and income-to-needs ratio. For each outcome, Model 2 considered outcomes with the interaction between race/ethnicity and income-to-needs ratio. To investigate the effects of each variable on outcomes within the race/ethnicity group, the analysis additionally conducted the logistic regression analysis, stratified by race/ethnicity.
**Bold-font:** Indicates statistically significant results.

Statistically significant results in Table 2, Model 1 show for DC, with all other covariates fixed, parents with a college degree or higher had lower DC (Odds Ratio (**OR**) 0.547, 95 percent [confidence interval (**95% CI**) 0.336 to 0.890] [*P*=0.015]) and higher income-to-needs (SES) levels with lower DC (OR 0.998, 95% [CI 0.997 to 0.999] [P=0.000]). For MOHP, Asian children had higher risk (OR 1.177, 95% [CI 0.839 to 1.651] [P=0.017]), parents with some college or higher education levels have lower risk (OR 0.527, 95% [CI 0.342 to 0.814] [P=0.004]), and higher income-to-needs (SES) levels with lower DC (OR 0.998, 95% [CI 0.997 to 0.999] [P=0.000]).

Statistically significant results in Table 3, Model 2 demonstrate for DC, lower risk is shown for Hispanic children have lower risk (OR 0.296, 95% [CI 0.185 to 0.474] [P=0.000]), parents with some college or higher (OR 0.553, 95% [CI 0.337 to 0.910] [P=0.020]), and higher income-to-needs (SES) levels (OR 0.998, 95% [CI 0.997 to 0.999] [P=0.000]). Also, for MOHP, higher risk is shown for Asian children (OR 2.497, 95% [CI 1.117 to 5.583] [P=0.026]), and with increasing levels of parental education. Children with higher income-to-needs (SES) levels had lower risk for MOHP (OR 0.998, 95% [CI 0.997 to 0.999] [P=0.000]). The most critical analysis for the effect of increased income-to-need ratio (SES) is shown in Table 3, Model 2 highlighted in gray color. Although all racial/ethnic groups benefitted equally from the protective effect of higher income and SES, for both DC and MOHP, no statistically significant differences were shown between racial/ethnic groups when all groups experienced increase income-to-needs ratio. Therefore, it might be construed that either MDR theory is not applicable to DC and MOHP or the biological disease processes for DC and MOHP supersede the health gain from higher SES.

## Discussion

### Findings and Hypothesis

The results of the current study, using 2017 NSCH data, failed to reject the null hypothesis and there is insufficient evidence to indicate a statistically significant difference between the racial/ethnic groups, with increased SES, and their clinical outcomes for dental caries and multiple oral health problems. This finding demonstrates that either 1) MDR theory is not applicable in the health outcomes of dental caries and multiple oral health problems for minority children, when comparing higher SES parity between White and minority populations or 2) the biological pathology of dental caries and multiple oral health problems have greater effect compared to the health gains from increased SES.

This present study’s findings showed no disparities between White and minority children for parental-reported DC and MOHP, even when all children gain higher SES.

### Comparison to Literature

The literature demonstrates strong support for MDR theory with evidence from a large number of studies focused on clinical conditions such as attention deficit hyperactive disorder, asthma, nutrition, smoking, drinking, obesity, impulse control, depression, anxiety, mental well-being, self-rated health, health care access, chronic diseases, mortality, and children’s dental access to and utilization of dental care.^16,19^ A New York Times published article, “Childbirth Is Deadlier for Black Families Even When They’re Rich, Expansive Study Finds,” which was based upon a study conducted by the National Bureau of Economic Research. The study found in the United States, the richest mothers and their newborns were the most likely to survive the year after childbirth, except when the family is Black. This groundbreaking study of two million California births supports MDR theory and showed the richest Black mothers and their babies were twice as likely to die as the richest White mothers and their babies.^20^

Hence, the finding that there is no difference in clinical outcomes between levels of family income is more likely because biologic effects causing dental caries may outweigh the protective factors of higher family income/SES. As described at a 2009 symposium sponsored by the American Dental Association, early childhood caries should be described as a “different” disease and instead be identified as a “pediatric infectious disease with dental manifestations, rather than simply a dental disease” due to the role of cariogenic bacteria, especially Streptococcus mutans (S. Mutans).^21^

Although there are many studies which associate higher caries prevalence with lower SES, ^22-28^ they do not specifically address the expected gain in health benefit from higher SES level from MDR theory. Hence, the findings from these studies imply that SES and education levels do not influence the rate of dental caries in children as much as other factors such as dietary habits and oral environment, such as the amount of S. mutans and biofilm, when trying to determine the relationship between caries prevalence and SES in a pediatric population.^29^

A body of epidemiologic studies supports biologic factors superseding MDR effects. Numerous epidemiologic studies provide evidence for the opposite effect with the association of higher SES levels with higher caries experience. One study by Psoter showed that family income levels were positively associated with any and maxillary incisor caries experience in pre-school children.^30^ Goodarzi reported that living in higher income households contributed as one of the factors contributing to higher DMFT scores in 12-year-old students in Tehran, Iran.^31^ Another study evaluating DMFT scores of children in Rome, Italy also highlights a positive relationship between DMFT scores and SES.^32^ More critically, a 2019 report by the Centers for Disease Control and Prevention (**CDC**) showed, from 1999-2004 and 2011-2016 National Health and Nutrition Examination Survey (NHANES) data cycles, the difference in primary/PERMANENT decayed and filled teeth (**dft/DFT**) between families with Federal Poverty Level (**FPL**) less than 100% and 100%-199%, regardless of race/ethnicity, was 1) zero percent for children aged two to eight years old; 2) an increase of five percent for children aged six to eleven years old; and 3) a small decrease of two percent for adolescents aged 12 to 19 years old.^25-28^ These data demonstrate that with increased family income, total lifetime tooth decay experience changed very little, if at all.

Clinical studies have also demonstrated strong evidence that biologic factors may influence the insidious nature of the dental caries process in all children. A 2019 study concluded that dental caries severity in the primary dentition was potentially related to individual genotypes and the level of oral *S. mutans* infectivity.^33^ In 2017 another study showed some children with severe early childhood caries (**S-ECC**) possessed a significantly different plaque microbiome comparted to caries-free children, with the S-ECC children experiencing higher levels of *S. mutans*.^34^ An interesting study demonstrated that the exponential expansion of *S. mutans* population of the oral cavity began approximately 10,000 years ago and coincided with the onset of human agriculture. This colonization pattern was associated with metabolic processes which contributed to the successful adaptation of *S. mutans* to its new niche, the human mouth, and with the dietary adaptations that coincided with the origin of agriculture.^35^ Hence, it may that humans are wedded to *S. mutans* and the resultant biologic effect of dental caries due to our dietary dependency on agriculture.

In sum, these epidemiological and clinical reports provide strong evidence that biologic factors influence the insidious nature of the dental caries process in all children and supersedes the effect of health gains from higher SES.

### Strengths and Limitations

A strength of this study is it is the first to apply MDR theory between multiple racial/ethnic groups for the outcomes of parental-reported DC and MOHP. In addition, the results of this study are generalizable with its large and diverse sample, high level of statistical power, and U.S. government data source which was nationally representative for children’s health.

However, the interpretation of this study must be carefully considered with appropriate limitations. This research study does not determine the cause-and-effect relationship between any variables used throughout the process of research. As well, the majority responders of the NSCH program were parents, and their interpretations of their children’s health might not be fully accurate. Since the entire data were extracted from a questionnaire-based survey, the question structures and word choices might affect the judgment of the responders which might also depend upon their family cultures and social experiences. Finally, the data were collected in 2017 and thus should be used with caution when applying the findings to current economic and health outcome conditions.

### Significance and Implications

It is important that dentists and policymakers realize that dental caries is an intractable disease that supersedes increased SES, and that clinical practice and government policies need to be made in the field of oral health education and prevention of caries to decrease the rate of childhood dental caries and oral health problems. Studies show that there is efficacy with implementing community fluoridated drinking water with a range of 30-60 percent less caries in fluoridated communities.^36^ Other studies show that the caries disease process becomes more prevalent when the oral environment’s pH decreases or when S mutans levels increase in the biofilm; caries prevalence may decrease with the addition of practices of increasing pH levels and decreasing biofilm and S. mutans levels practices (such as by rinsing the mouth with baking soda or chewing xylitol gum after eating) to one’s daily oral hygiene maintenance.^28^ Future policies regarding pediatric oral health care should address the above-noted factors contributing to increased rates of dental caries and oral health problems through increasing oral health literacy for both parents and their kids, emphasizing the importance of and offering incentives for regular, periodic dental visits for maintenance of oral health. More nutritional counseling and proper oral hygiene care education should be provided by dental practitioners. More policies should be enacted that contribute to prevention of dental caries such as subsidizing the cost of promoting the use of xylitol as a sweetener and providing fluoridated water in every community. More mandatory continuing education courses on the topic of caries prevention and oral hygiene education can be provided to dentists to remind and teach their patients at every visit. Oral health clinicians should also be aware that although MDR theory was not directly applicable to pediatric caries rates, minority groups of children still endure oral disparities through systemic and structural racism prevalent in society and solving those issues will help with increase in access and utilization of dental care to be able to then treat caries in the pediatric population.

### Future Studies

To strengthen the evidence for MDR theory, future studies might be designed with prospective and longitudinal constructs which generate predictive statistical analyses. This study’s findings, generated by the 2017 NSCH data, indicated MDR theory cannot be applied to childhood caries rates. Hence, future studies might also explore more recent NSCH data sets to learn what types of policies enacted may help decrease the rate of childhood caries.

## Conclusion

Based on this study’s results, the following conclusions can be made:

1. Among all racial/ethnic groups and different levels of SES of children, Minorities’ Diminished Return theory was not applicable to the outcomes of childhood caries and multiple oral health problems.
2. The biologic disease process for dental caries and multiple oral health problems may supersede the effect of Minorities’ Diminished Return theory.
3. Dentists and policymakers must support policies and best practices which benefit entire populations of children, through widespread prevention, access-to-care, and utilization-of-care programs to decrease levels of childhood caries and other oral health problems.

## Data Availability

All data produced in the present study are available upon reasonable request to the authors

## Acknowledgments

The authors thank the Hansjörg Wyss Department of Plastic Surgery, NYU Langone Health for funding; Veronica Brandley, DMD, Paulina Nguyen, DDS, and Regina Nguyen, DDS who assisted this project during residency training; and Jay Balzer, DMD, MPH, Amy Kim, DDS, and Adolfina Polk, DDS who provided manuscript review.

## Reference

1. AMA Board of Trustees pledges action against racism, police brutality. Press Release. American Medical Association. Published, June 07, 2020. Accessed November 10, 2022. https://www.ama-assn.org/press-center/press-releases/ama-board-trustees-pledges-action-against-racism-police-brutality

2. Feagin JR, Ducey K. Racist America: Roots, Current Realities, and Future Reparations. 4th ed. Routledge; 2018. Accessed May 15, 2022. https://doi.org/10.4324/9781315143460

3. Bonilla-Silva E. Rethinking racism: Toward a structural interpretation. American Sociological Review; 62(3):465–480. https://doi.org/10.2307/2657316

4. Flores G, Lin H. Trends in racial/ethnic disparities in medical and oral health, access to care, and use of services in US children: has anything changed over the years? Int J Equity Health. 2013;12(1):10. doi:10.1186/1475-9276-12-10

5. Flores G, Tomany-Korman SC. Racial and ethnic disparities in medical and dental health, access to care, and use of services in US children [published correction appears in Pediatrics. 2009 Sep;124(3):999-1000]. Pediatrics. 2008;121(2):e286–e298. doi:10.1542/peds.2007-1243

6. Case A, Lubotsky D, Paxson C. Economic status and health in childhood: The origins of the gradient. The American Economic Review. 2002;92(5):71.

7. Akinbami LJ, Moorman JE, Simon AE, Schoendorf KC. Trends in racial disparities for asthma outcomes among children 0 to 17 years, 2001-2010. J Allergy Clin Immunol. 2014;134(3):547-553.e5. doi:10.1016/j.jaci.2014.05.037

8. Minnick ML, Boynton S, Ndirangu J, Furth S. Sex, race, and socioeconomic disparities in kidney disease in children. Semin Nephro. 2010;30(1):26–32. doi:10.1016/j.semnephrol.2009.10.003

9. Como DH, Stein Duker LI, Polido JC, Cermak SA. The persistence of oral health disparities for African American children: a scoping review. Int J Environ Res Public Health. 2019;16(5):710. doi:10.3390/ijerph16050710

10. Chaffee BW, Rodrigues PH, Kramer PF, Vítolo MR, Feldens CA. Oral health-related quality of life measures: variation by socioeconomic status and caries experience. Community Dent Oral Epidemiol. 2017;45(3):216–224. doi:10.1111/cdoe.12279

11. Kelesidis N. A racial comparison of sociocultural factors and oral health perceptions. J Dent Hyg. 2014;88(3):173–182.

12. Assari S, Hani N. Household income and children’s unmet dental care need; Blacks’ diminished return. Dent J (Basel). 2018;6(2):17. doi:10.3390/dj6020017

13. Assari S. Health disparities due to diminished return among Black Americans: public policy solutions. Soc Issues Policy Rev. 2018;12(1):112–145. doi:10.1111/sipr.12042

14. Assari S. Unequal gain of equal resources across racial groups. Int J Health Policy Manag. 2017;7(1):1–9. doi:10.15171/ijhpm.2017.90

15. Assari S, Moghani Lankarani M. Poverty status and childhood asthma in White and Black families: National Survey of Children’s Health. Healthcare (Basel). 2018;6(2):62. doi:10.3390/healthcare6020062

16. Assari S. Special Issue “Health disparities due to minorities’ diminished returns (MDRs).” Int J Environ Res Public Health. 2019. Accessed November 10, 2022. https://www.mdpi.com/journal/ijerph/special_issues/MDRs

17. National Survey of Children’s Health. NSCH codebooks. NSCH Codebooks - Data Resource Center for Child and Adolescent Health. Published 2019. Accessed November 10, 2022. https://www.childhealthdata.org/learn-about-the-nsch/nsch-codebooks

18. R Core Team (2018). R: A language and environment for statistical computing. R Foundation for Statistical Computing, Vienna, Austria. Published 2018. Accessed November 10, 2022. https://www.R-project.org/

19. Okuji, D, Wei T, Lee M. Family Income and Utilization Disparities for Dental Access of Minority Children: A Cross-Sectional Study. medRxiv 2023.02.07.23285585; doi: https://doi.org/10.1101/2023.02.07.23285585

20. Kennedy-Moulton K, Miller S, Miller, Persson P, Rossin-Slater M, Wherry L, Aldana G. Maternal and Infant Health Inequality: New Evidence from Linked Administrative Data. National Bureau of Economic Research Working Paper Series. 2022(Series # 30693). Accessed April 14, 2023. https://www.nber.org/system/files/working_papers/w30693/w30693.pdf

21. Symposium on Early Childhood Caries in American Indian and Alaska Native Children Council on Access, Prevention and Interprofessional Relations Panel Report. Updated November 2009. Accessed April 14, 2023.https://www.ada.org/-/media/project/ada-organization/ada/ada-org/files/resources/research/oral-health-topics/topics_caries_symposium.pdf?rev=7014596d6a764da68aa740985b6c41bc&hash=AEC725EB27BA9BA1A36C1CD244C7DE0F

22. Popoola, B. O., Denloye, O. O., & Iyun, O. I. (2014, April 4). Influence of parental socioeconomic status on caries prevalence among children seen at the University College Hospital, Ibadan. Annals of Ibadan Postgraduate Medicine. Retrieved December 30, 2022. https://www.ajol.info/index.php/aipm/article/view/102555

23. Chen, K. J., Gao, S. S., Duangthip, D., Li, S. K., Lo, E. C., & Chu, C. H. (2017). Dental caries status and its associated factors among 5-year-old Hong Kong Children: A cross-sectional study. BMC Oral Health, 17(1). https://doi.org/10.1186/s12903-017-0413-2

24. Nomura L., Bastos, J., Peres M. (2002). Dental pain prevalence and association with dental caries and socioeconomic status in schoolchildren. Scientific Electronic Library Online. Retrieved December 31, 2022. https://www.scielo.br/j/bor/a/rdS8bFYGwDCzHpFWQTFLL6d/abstract/?lang=en

25. Table 3. Centers for Disease Control and Prevention. Oral Health Surveillance Report: Trends in Dental Caries and Sealants, Tooth Retention, and Edentulism, United States, 1999–2004 to 2011–2016. Atlanta, GA: Centers for Disease Control and Prevention, U.S. Department of Health and Human Services; 2019. Accessed April 14, 2023. https://www.cdc.gov/oralhealth/publications/OHSR2019-table-3.html

26. Table 7. Centers for Disease Control and Prevention. Oral Health Surveillance Report: Trends in Dental Caries and Sealants, Tooth Retention, and Edentulism, United States, 1999–2004 to 2011–2016. Atlanta, GA: Centers for Disease Control and Prevention, U.S. Department of Health and Human Services; 2019. Accessed April 14, 2023. https://www.cdc.gov/oralhealth/publications/OHSR2019-table-7.html

27. Table 11. Centers for Disease Control and Prevention. Oral Health Surveillance Report: Trends in Dental Caries and Sealants, Tooth Retention, and Edentulism, United States, 1999–2004 to 2011–2016. Atlanta, GA: Centers for Disease Control and Prevention, U.S. Department of Health and Human Services; 2019. Accessed April 14, 2023. https://www.cdc.gov/oralhealth/publications/OHSR2019-table-11.html

28. Table 16. Centers for Disease Control and Prevention. Oral Health Surveillance Report: Trends in Dental Caries and Sealants, Tooth Retention, and Edentulism, United States, 1999–2004 to 2011–2016. Atlanta, GA: Centers for Disease Control and Prevention, U.S. Department of Health and Human Services; 2019. Accessed April 14, 2023. https://www.cdc.gov/oralhealth/publications/OHSR2019-table-16.html

29. Zou J, Du Q, Ge L, et al. Expert consensus on early childhood caries management. Int J Oral Sci. 2022;14(1):35. Published 2022 Jul 14. doi:10.1038/s41368-022-00186-0

30. Psoter WJ, Pendrys, DG, Morse, DE, Zhang, H, & Mayne, ST. Associations of ethnicity/race and socioeconomic status with early childhood caries patterns. Journal of Public Health Dentistry. 2006;66(1), 23–29. https://doi.org/10.1111/j.1752-7325.2006.tb02547.x

31. Goodarzi A, Heidarnia A, Tavafian SS, Eslami M. Evaluation of Decayed, Missing and Filled Teeth (DMFT) Index in the 12 Years Old Students of Tehran City, Iran. Brazillian Journal of Oral Sciences. September, 21, 2021. Accessed April 14, 2023. https://pdfs.semanticscholar.org/b961/ab95c7dc54de45c68d191d4685cb383346ba.pdf

32. Costacurta M, Epis M, Docimo R. Evaluation of DMFT in paediatric patients with social vulnerability conditions. Eur J Paediatr Dent. 2020;21(1):70–73. doi:10.23804/ejpd.2020.21.01.14

33. Meng Y, Wu T, Billings R, Kopycka-Kedzierawski DT, Xiao J. Human genes influence the interaction between Streptococcus mutans and host caries susceptibility: a genome-wide association study in children with primary dentition. Int J Oral Sci. 2019;11(2):19. Published 2019 May 30. doi:10.1038/s41368-019-0051-4

34. Agnello M, Marques J, Cen L, et al. Microbiome Associated with Severe Caries in Canadian First Nations Children. J Dent Res. 2017;96(12):1378–1385. doi:10.1177/0022034517718819

35. Cornejo OE, Lefébure T, Bitar PD, et al. Evolutionary and population genomics of the cavity causing bacteria Streptococcus mutans. Mol Biol Evol. 2013;30(4):881–893. doi:10.1093/molbev/mss278

36. Newbrun, E. (1989). Effectiveness of water fluoridation. Journal of Public Health Dentistry. 1989;49(5), 279–289. https://doi.org/10.1111/j.1752-7325.1989.tb02086.x

